# An *“In-House”* ELISA for SARS-CoV-2 RBD uncovers elevated immune response at higher altitudes

**DOI:** 10.1101/2021.03.10.21252711

**Authors:** Tomas Grau Rodrigo, Ploper Diego, Ávila César, Vera Pingitore Esteban, Maldonado Carolina, Chaves Silvina, Socias Sergio Benjamín, Stagnetto Agustín, Navarro Silvia, Chahla Rossana, Aguilar Mónica, Llapur Conrado, Aznar Patricia, Alcorta Malena, Costas Dardo, Flores Isolina, Heinze Dar, Apfelbaum Gabriela, Mostoslavsky Raúl, Mostoslavsky Gustavo, Cazorla Silvia, Perdigón Gabriela, Chehín Rosana

## Abstract

The severe acute respiratory syndrome coronavirus-2 (SARS-CoV-2) first reported in Wuhan has caused a global pandemic with dramatic health and socioeconomic consequences. The Coronavirus Disease 2019 (COVID-19) associated represents a challenge for health systems that had to quickly respond developing new diagnostic and therapeutic strategies. In the present work, we developed an “*In House*” ELISA with high sensitivity (92.2 %), specificity (100%) and precision (93.9%), with an area under the ROC curve (AUC) of 0.991, rendering the assay as an excellent serological test to correctly discriminate between SARS-COv-2 infected and non-infected individuals and study population seroprevalence. Among 758 patients evaluated for SARS-CoV-2 diagnosis in the province of Tucumán, Argentina, we found a Pearson correlation coefficient of 0.5048 between antibodies elicited against the RBD and the nucleocapsid (N) antigen. Additionally, 33.6% of individuals diagnosed with COVID-19 displayed mild levels of RBD-IgG antibodies, while 19% of the patients showed high antibody titers. Interestingly, patients with SARS-COV-2 infection over 60 years old elicited significantly higher levels of IgG antibodies against RBD compared to younger ones, while no difference was found between women and men. Surprisingly, individuals from a high altitude village displayed statistically significant higher and longer lasting anti-RBD antibodies compared to those from a city at a lower altitude, suggesting that a hypobaric hypoxia-adapted mechanism may act as a protective factor for COVID-19. To our knowledge, this is the first report correlating altitude with increased humoral immune response against SARS-Cov-2 infection.

## Introduction

The severe acute respiratory syndrome coronavirus 2 (SARS-CoV-2) virus that emerged in December 2019, with its associated Coronavirus Disease (COVID-19), has caused more than 2 million deaths in 2020 alone ^1^. It is projected that without efficient action, COVID-19 could infect around 90% of the world’s population and kill over 40 million people. While vaccination efforts continue, the near future holds no end in sight for this pandemic ^2^. Diagnostic efficiency continues to be one of the main bottlenecks in the fight against COVID-19. While nucleic acid detection of the SARS-CoV-2 RNA genome is the gold standard test to diagnose infection, measurements of antibody responses give essential knowledge regarding protective immunity due to infection or vaccination. Serological surveillance of anti-SARS-CoV-2 antibodies in the population also provides a crucial tool for designing public health guidelines^3^. This is only possible if serological tests are sufficiently trustworthy, which basically implies the correct election of the technique, the target antigen, and the antibodies to be studied.

The SARS-CoV-2 pandemic has disrupted the worldwide supply chain for many diagnostic equipment and their components, challenging government agencies and private companies in their efforts to acquire reagents, testing kits, personal protective equipment, laboratory devices, vaccines, and any COVID-19-related technology. While this situation has provided fertile ground for the development of new private or state-owned companies that have flourished attempting to resolve this urgent demand, supply is still proving insufficient. This is also the case for kits that measure antibodies against SARS-CoV-2. Therefore, in order to resolve this shortage and support state-led diagnostic efforts, many research laboratories have repurposed their expertise, skilled labor and equipment by developing and assembling *in-house* testing kits, among other efforts ^4-7^.

SARS-CoV-2 serological tests employ enzyme-linked immunosorbent assay (ELISA)^8,9^, immunofluorescence^10^ and lateral flow techniques to detect antibodies directed against the nucleocapsid (N) and/or Spike (S) proteins of the viral proteome^11^. The highly glycosylated S protein, which plays a key role in the receptor recognition and cell membrane fusion process, is composed of two subunits, S1 and S2. The S1 subunit contains a receptor-binding domain (RBD) that recognizes and binds to the host receptor angiotensin-converting enzyme 2 (ACE2)^12,13^. The N protein plays a vital role in transcription and replication and, due to its abundance, has been suggested to be the ideal antigen for detecting early infections ^3^. It has been shown that IgG antibodies targeting S are more specific, while those directed against N may be more abundant in the earliest phase of infection ^14,15^. The sensitivity and specificity of most serological tests have been validated with samples extracted from patients enduring the acute phase of infection ^14,15^. Results from these studies indicate that tests using N and S were considered equally sensitive ^14,15^. Based on these observations, it has been assumed that determination of antibody responses against either protein would be equally suitable in post-infection phase population-based seroprevalence studies^16^. However anti-S and anti-RBD antibodies correlate better with virus neutralization ^17-20^. This has been determined through virus neutralization tests (VNT) (the gold standard for measuring neutralization). Since this assay is labor-intensive and requires a highly trained staff working in BSL3 conditions, it is not appropriate for high-throughput detections. In contrast, ELISA with recombinant S and/or RBD as substrate, show a strong correlation with neutralization assay results ^20,21^. High-titer plasma therapy is one of the few clinically proven treatments that prevent COVID-19 in adults. Thus, determining the levels of neutralizing antibodies is fundamental when screening samples for potential plasma donors ^22^.

In addition, since most of the vaccines approved to date carry the genetic information of S or its RBD domain, only tests that determine anti-S or anti-RBD antibodies can evaluate individual and population seroconversions triggered by vaccination ^23,24^. The use of RBD not only streamlined the diagnostic kit production process but also provides a more specific target for neutralizing antibodies^25,26^. Therefore, determining the titers of anti-S or anti-RBD antibodies, instead of anti-N, provides more valuable information.

Taking into context the worldwide demand for antibody testing kits and the importance of measuring the seroconversion in the population, the aim of the present study was to develop an “*In House*” ELISA to detect anti-RBD antibodies in the SARS-CoV-2 convalescent population from Tucumán-Argentina, in order to contribute to local public health policies. The particular geographical location of Tucumán in the Andean foothills allowed us to compare the anti-RBD titers developed by individuals in low altitude locations in contrast to a high-altitude hypobaric hypoxia-adapted population. We herein report that inhabitants of Tafi del Valle, a village situated at 2014 meters above median sea level (mamsl), showed statistically significant higher anti-RBD titers than the population of San Miguel de Tucumán, located at 431 mamsl. To our knowledge, this is the first study showing an increase in the humoral immune response against SARS-CoV-2 in a high-altitude population, providing a novel perspective on the hypothesis of increased resistance to COVID-19 in populations adapted to hypobaric hypoxia^27-29^.

## Materials and Methods

### Recombinant SARS-CoV-2 S RBD-His expression

A plasmid encoding a secreted his-tagged SARS-CoV-2 S RBD was obtained as a generous gift from Jared Feldman and Aaron G. Schmidt (Harvard University). The RBD sequence was PCR amplified and subcloned into a pHAGE2-EF1a-IRES-ZsGreen-W lentiviral vector^30^. Lentiviral particles were produced by cotransfecting the lentiviral backbone together with appropriate packaging vectors into HEK293 cells at 70% confluency in a 100 mm petri dish. HEK293 were grown in high glucose medium supplemented with 10% fetal bovine serum and antibiotic/antimycotic and PEI 87 kDa was used as the transfection reagent. Culture media containing lentiviral particles was harvested after 24/48 h and used to transduce a fresh HEK293 culture at 70% confluency in one well of a 6 well plate. Media was washed after 24 hs. Transduction efficiency was assessed by imaging the fluorescence of ZSGreen, expressed from the pHAGE2 lentiviral vector.

### Recombinant SARS-CoV-2 S RBD-His purificatio

Purification of each RBD-His contained in the conditioned media was carried out by affinity chromatography using a 5 ml HisTrap HP column (GE Healthcare, UK) coupled to an Akta Pure 20 l (GE) chromatograph (FPLC). Briefly, the supernatant was harvested from the cell culture, supplemented with 20 mM imidazole and filtered through a 0.22 um pore size PVDF filter. The affinity column was equilibrated with 10 column volumes of buffer A (20 mM NaH2PO4, NaCl 500 mM, pH 7.4, 20 mM imidazole), loaded with 100 ml sample using a sample pump, and washed with 10 column volumes of buffer A at a flow rate of 4 ml/min. Bound proteins were eluted with a step gradient with 5 CV of either 20%, 50% and 100% of buffer B (20 mM NaH2PO4, NaCl 500 mM, pH 7.4, 500 mM imidazole). Fractions of 5 ml were collected and further analyzed using SDS-PAGE. Fractions with molecular weight compatible with the His-tagged RBD were pooled and dialyzed against 200 mM NaH2PO4, pH 6.5. The production yield for RBD was around 0.5 mg/100 ml of cell culture supernatant, with a purity level higher than 95%.

### ELISA anti-RBD antibody binding assay

An indirect ELISA test for the determination of IgG anti–RBD antibody was design. Briefly, flat polystyrene bottom plates (High Binding, Half-Area, Greiner 675061) were sensitized with 0.1 µg per well of the RBD antigen for 18 h at 4°C. Blocking was done with 10% Fetal Bovine Serum (FBS) in PBS 10mM pH7.4 during 1 h at 37 °C. Plates were then washed three times with 0.1% Tween in PBS. Sera were assayed at a serial dilution of 1/100 and incubated for 1 h at 37°C. Peroxidase-conjugated immunoglobulins to human IgG (whole molecule) -Peroxidase (Sigma A8667) diluted 1/35,000 were used as a secondary antibody. Plates were developed by adding TMB (3,3’,5,5’ – Tetrametilbenzidina; BD OptEIAtm), incubated for 15 min in the dark and the reaction was stopped using 4N H2SO4. Optical density was read by an ELISA reader (TECAN Spark) at 450 nm. Cutoff values were calculated using receiver operating characteristic curve (ROC). Titers were calculated as the dilution in which the optical density (OD) obtained was equal to the cutoff.

### Populations studied

Serum samples (758) taken in the Laboratorio de Salud Pública-Sistema Provincial de Salud de Tucumán (SIPROSA) from patients with diagnosis of SARS-Cov-2 infection by positivity results in the polymerase chain reaction (PCR) or due to close contact to SARS-Cov-2 infected peoples were analyzed. Serum samples were taken at least 3 weeks after the diagnosis of Covid-19. The procedures were approved by the Bioethics Committee of the SIPROSA N° 29/2020. As control of no SARS-Cov-2 infection 26 serum taken before December of 2019 were analyzed (prepandemic serum). RBD specific IgG levels along time was evaluated in 574 and 14 patients, from a high-altitude village (Tafí del Valle, population 14933, representing 0.093% of the population), and a lower altitude city (San Miguel de Tucumán, population 830000, representing 0.069% of the population), respectively, with SARS-CoV-2 infection confirmed by RT-PCR.

### Statistical analyses

The cutoff point for optimal sensitivity and specificity, as well as the other statistical parameters, were determined using the receiver operating characteristic (ROC) curve analysis using the XL-STAT statistical software/program (Excel). RBD-IgG titers, and correlation and differences between medias studies were carried out with the Prisma 8.0 Software (GraphPad, San Diego, CA). For non-parametric variables, data were analysed by the Kruskal-Wallis test. Significant differences between groups are shown with the corresponding P values. Significant differences are indicated with asterisks.

## Results

### 1) Expression and purification of the recombinant RBD-SARS-CoV-2

In order to evaluate local population anti-SARS-CoV-2 IgG antibody levels, an “*In House*” ELISA assay for SARS-CoV-2 which uses the RBD of S was developed. For this, a transgenic HEK293 cell line was produced that expresses and secretes the His-tagged RBD (HEK293-RBD-His-SARS-CoV-2). This stable cell line, which secretes large amounts of the RBD-His into the culture medium, was achieved by transducing HEK293 cells with a pHAGE2 lentivirus (pHAGE-RBD-His), generated by cotransfecting HEK293 cells with the appropriate lentiviral vector and corresponding packaging plasmids (*materials and methods*) (Fig. 1A-C). The secreted RBD-His domain, efficiently purified by affinity chromatography, migrated as expected on an SDS-PAGE gel according to previous reports (Fig. 1D-E) ^31^.

**Figure 1.**
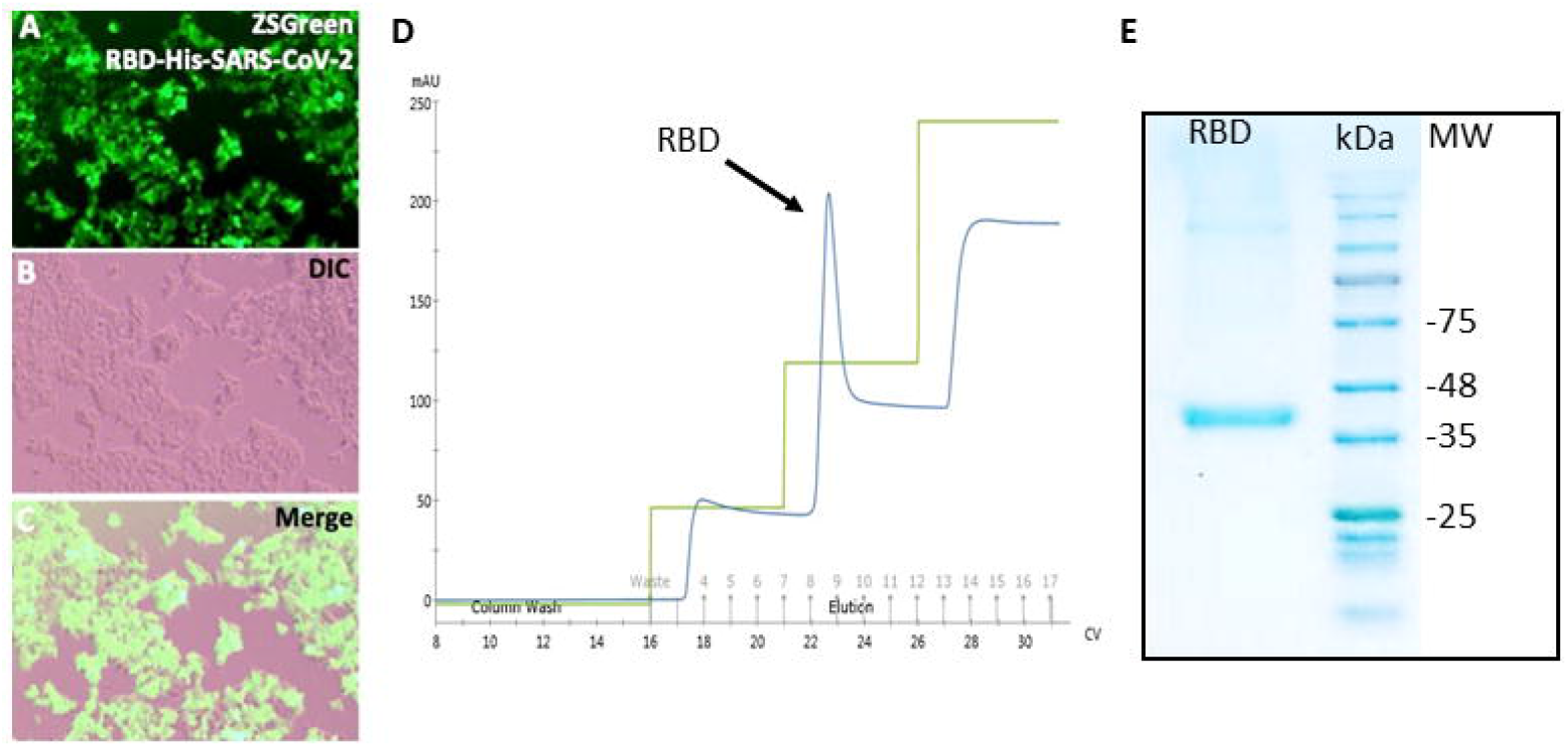
Expression and purification of the recombinant RBD-SARS-CoV-2. **(A-C)** Highly efficient transduction of HEK293 cells with pHAGE2 lentiviral particles coding for a secreted form of RBD-His can be observed by expression of the fluorescent reporter ZsGreen, co-expressed from the same lentivirus. **D)** Elution profile of the supernatant from the stable transgenic cell line HEK293-RBD-His-SARS-CoV-2 produced. The purified protein appeared in the second absorbance peak (blue) as indicated (arrow). **E**) SDS-PAGE of RBD-His purified by an HPLC system using HisTrap columns. Lane 1: purified RBD showing the expected molecular weight; lane 2: molecular weight marker.

### 2) Diagnostic performance of an “*In House*” ELISA using purified SARS-CoV-2 RBD

A titration curve was performed to determine the most appropriate concentration of the RBD antigen to be used in the ELISA (data not shown). Then, ELISA were performed to determine the titer of specific IgG antibodies against RBD in serum samples from convalescent serum of patients of the local state health system (SIPROSA-Tucumán, Argentina). As positive controls for SARS-CoV-2 infection, we analysed 52 patients that presented positive results both in the PCR test and in the Chemiluminescent microparticle immunoassay (CMIA-Architect, Abbot) performed on the nasopharyngeal swab and on serum samples, respectively. As shown in Fig. 2A, anti-RBD IgG antibody levels were significantly higher in samples from these patients previously diagnosed with SARS-CoV-2 than in serum samples extracted from pre-pandemic patients (p< 0.0001) (Fig. 2A). Next, we analysed the accuracy of our ELISA test to correctly classify samples as positive for SARS-CoV-2 infection. As shown in Fig. 2B, the AUC 0.991 (95% confidence interval 0.9778 to 1.000) was determined for the RBD antigen. Moreover, the ELISA for the determination of IgG anti-RBD presented high sensitivity (92.2%), specificity (100%) and also a predictive positive value (1.00) (Fig. 2C). In order to determine if antibodies present in serum elicited by other microorganisms can interfere with the ELISA test, sera extracted from patients before December 2019 with human immunodeficiency virus (HIV), *Toxoplasma gondii, Trypanosoma cruzi*, and human hepatitis A virus (HAV) infections were tested. As shown in Fig. 2D, no false positive results arose from these samples (**** p<0.0001).

**Figure 2.**
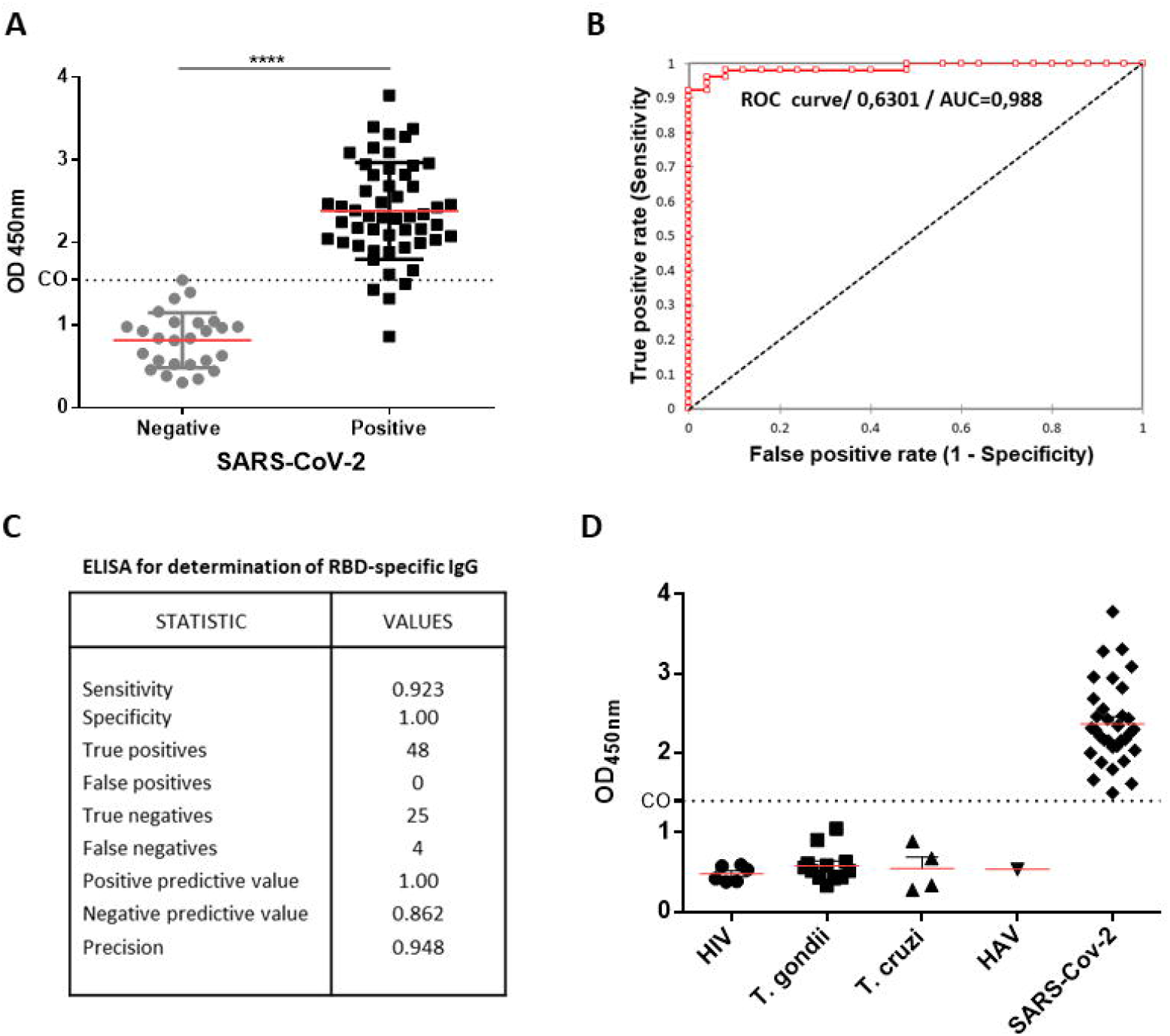
Diagnostic performance of an “*In House*” ELISA using purified SARS-CoV-2 RBD. **A)** ELISA test for IgG antibodies against RBD. Serum samples were classified as positive for SARS-CoV-2 infection by being positive in the PCR and the CMIA test. Negative samples for SARS-CoV-2 infection were serum taken before December 2019. The results were expressed as the OD450 nm, and the cutoff (CO) was calculated using the ROC curve. **B)** Diagnostic efficacy of the RBD antigens in SARS-CoV-2 infection using ROC curves. **C)** Statistic parameters of the ELISA test developed against the RBD antigen. **D)** IgG antibodies against RBD in sera from patients with infections by: HIV: human immunodeficiency virus; T. gondii: *Toxoplasma gondii*; T. cruzi: *Trypanosoma cruzi*; and HAV: human hepatitis a virus (**** p<0.0001).

### 3) Performance of an “*In House*” ELISA compared to a commercially available test

The ability to detect antibodies triggered by SARS-CoV-2 infection was compared between the anti-RBD ELISA and a commercially available Chemiluminescent Microparticle ImmunoAssay (CMIA) kit that detects antibodies against the N protein (Architect, Abbott). Among 595 patients that were classified as SARS-CoV-2-positive by RT-PCR from the local public health system, we observed that the ELISA and the CMIA-based Architect displayed similar diagnostic abilities, 59% and 58.3%, respectively (Fig. 3A). Interestingly, similar results were observed in patients referred to as close contacts to SARS-CoV-2 infected patients, as both tests were able to detect antibodies in similar rates within this population (anti-RBD ELISA, 63.8%; anti-N CMIA, 58.9 %) (Fig. 3A).

**Figure 3.**
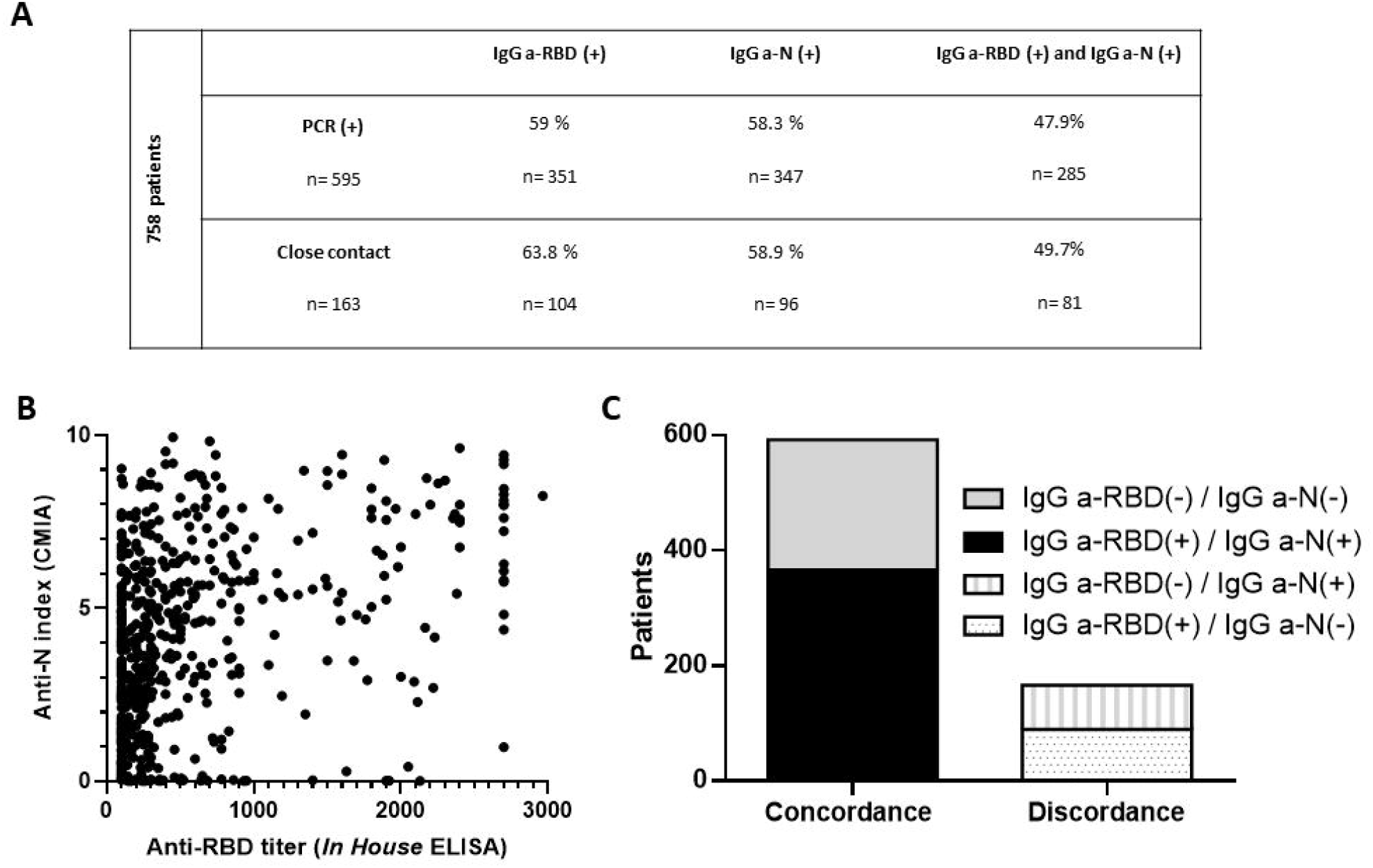
Performance of the anti-RBD “*In House*” ELISA compared to a commercially available test. **A)** Percentages of the population studied (n=758), either diagnosed as SARS-CoV-2 positive by RT-PCR or close contacts of infected patients, that have detectable SARS-CoV-2 anti-RBD or anti-N antibodies as measured by the “*In House*” ELISA or CMIA, respectively. **B)** Scatterplot depicting the relationship between the titers from the anti-RBD ELISA and the anti-N CMIA index for 758 patients. **C)** Concordance or discordance in results from the anti-RBD ELISA and the anti-N CMIA tests in the screening of IgG antibodies elicited after SARS-CoV-2 infection.

We further analysed the correlation between both tests, and found a Pearson correlation coefficient of 0.5048 between anti-RBD IgG titers and the anti-N IgG index (Fig. 3B). Albeit this weak correlation between the titers of anti-RBD and the CMIA index of anti-N antibodies, a high concordance for the presence or absence of both antibodies was observed (Fig 3C). Therefore, as shown in Fig. 3B, high anti-N IgG antibodies in a given patient does not necessarily reflect stimulation of high humoral immune response against RBD. This fact could be due to differences in antigen expression levels or time elapsed after infection. Overall, these results endorse the “*In House*” ELISA for the detection of anti-RBD IgG as an excellent candidate for the determination of humoral immune responses elicited against SARS-Cov-2 infection.

### 4) Distribution of anti-RBD specific IgG antibodies measured with an “*In House*” ELISA among patients diagnosed with SARS-CoV-2

In order to study the distribution of anti-RBD IgG titers among true positive samples, we analysed, with the “*In House*” ELISA, the sera of 366 patients previously diagnosed with SARS-CoV-2 by RT-PCR and who also tested positive for anti-N antibodies by CMIA.

The distribution of antibody levels showed a preponderance of titers between 300 and 800 (37%), where 19% of titers are above 1350 (Fig. 4A). When 758 patients that tested positive for CMIA anti-N were analysed by the “*In House*” ELISA and segregated into three arbitrary age groups, ?40 years, 40-60 years and <60 years, we found that IgG anti-RBD antibodies were significantly elevated in the over-60 age group, as previously reported ^32^. No significant differences were found in average RBD-specific IgG titers between male and female patients (Fig. 4C), although the percentage of negativity was much higher in males (48.9%) than in females (9.7%) in the studied population with the anti-RBD ELISA. Overall, the distribution of RBD-specific IgG antibodies in the population of Tucumán was found to be consistent with what has been described in previous seroprevalence reports^33,34^

**Figure 4.**
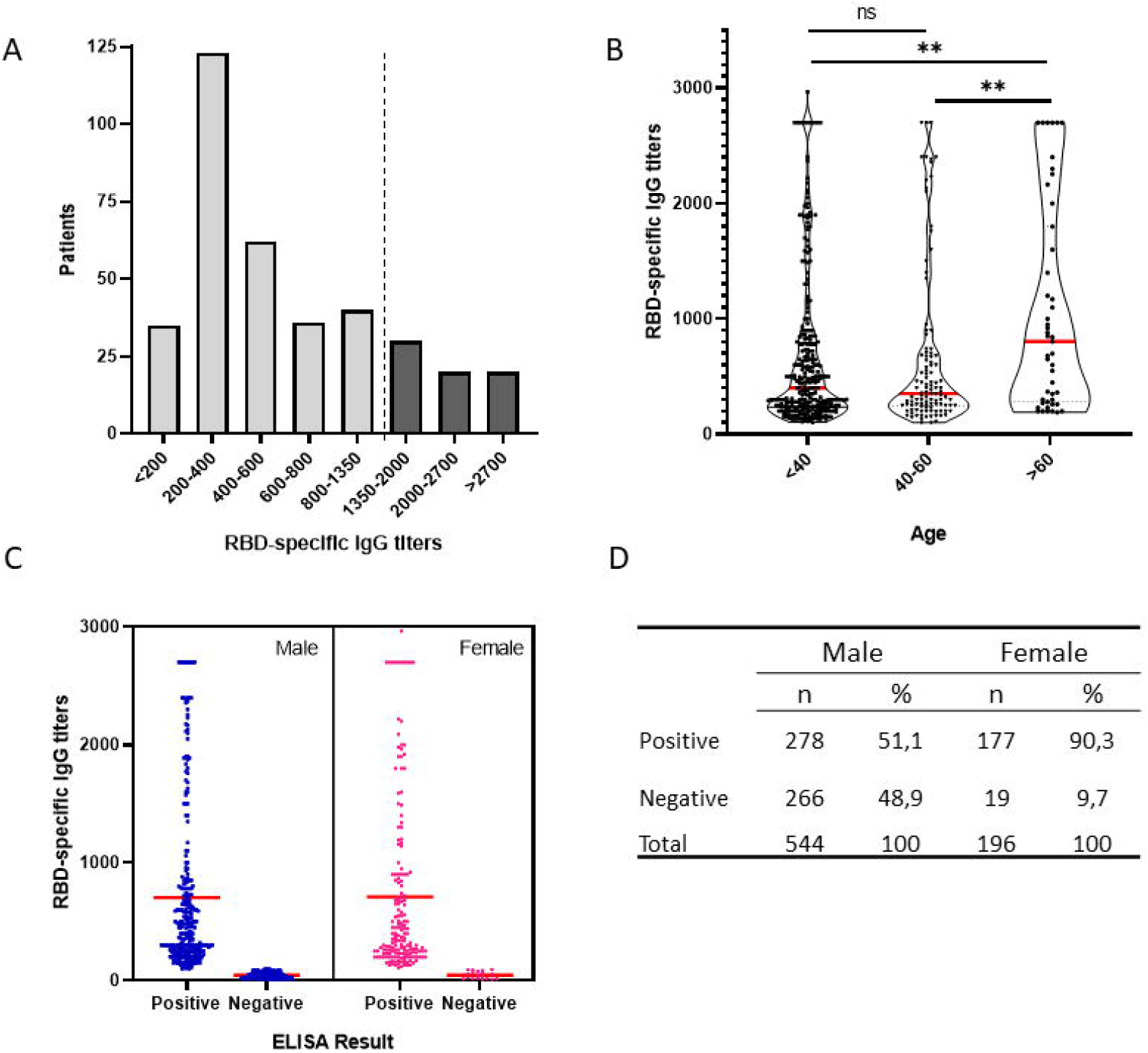
IgG anti-RBD antibody titers among patients measured by an “*In House*” ELISA. A) Distribution of anti-RBD titers among 366 true positive samples. Dot line delimits the population that correlates to a probability ≥80% of having neutralizing titers ≥160^21^. B) Distribution of anti-RBD titers of 758 patients segregated into three age groups, patients under 40 (<40), 40-60, and above 60 (>60) years old. C) Distribution of anti-RBD titers according to sex. D) Chart detailing the number (n) and percentage (%) of male or female patients who tested positive or negative for the presence of anti-RBD antibodies in an “*In House*” ELISA.

### 5) Increased and long-lasting anti-RBD humoral immune response in a high altitude population

In addition to serum samples corresponding to patients from the city of San Miguel de Tucumán (431 mamsl), we also measured anti-RBD IgG titers in patients from Tafí del Valle, a small village located at 2014 mamsl. Surprisingly, we observed significantly higher anti-RBD IgG antibody levels in patients from this high altitude village compared to the lower altitude population (p<0.001) (Fig. 5A). Interestingly, high altitude patients sustained specific antibody titers for extended periods of time, showing similar IgG specific antibodies levels at day 90 and 30 post COVID-19. By contrast IgG anti-RBD IgG antibodies significantly decrease at day 90 post-diagnosis of viral infection with respect to the levels observed at day 30 in individuals from the lower altitude population studied. The fact that this ratio of antibodies between day 90 and 30 in these two populations show significant differences (p<0.005) (Fig. 5B), suggests that a hypobaric hypoxia-adapted mechanism might play a key role in the SARS-CoV-2 humoral immune response.

**Figure 5.**
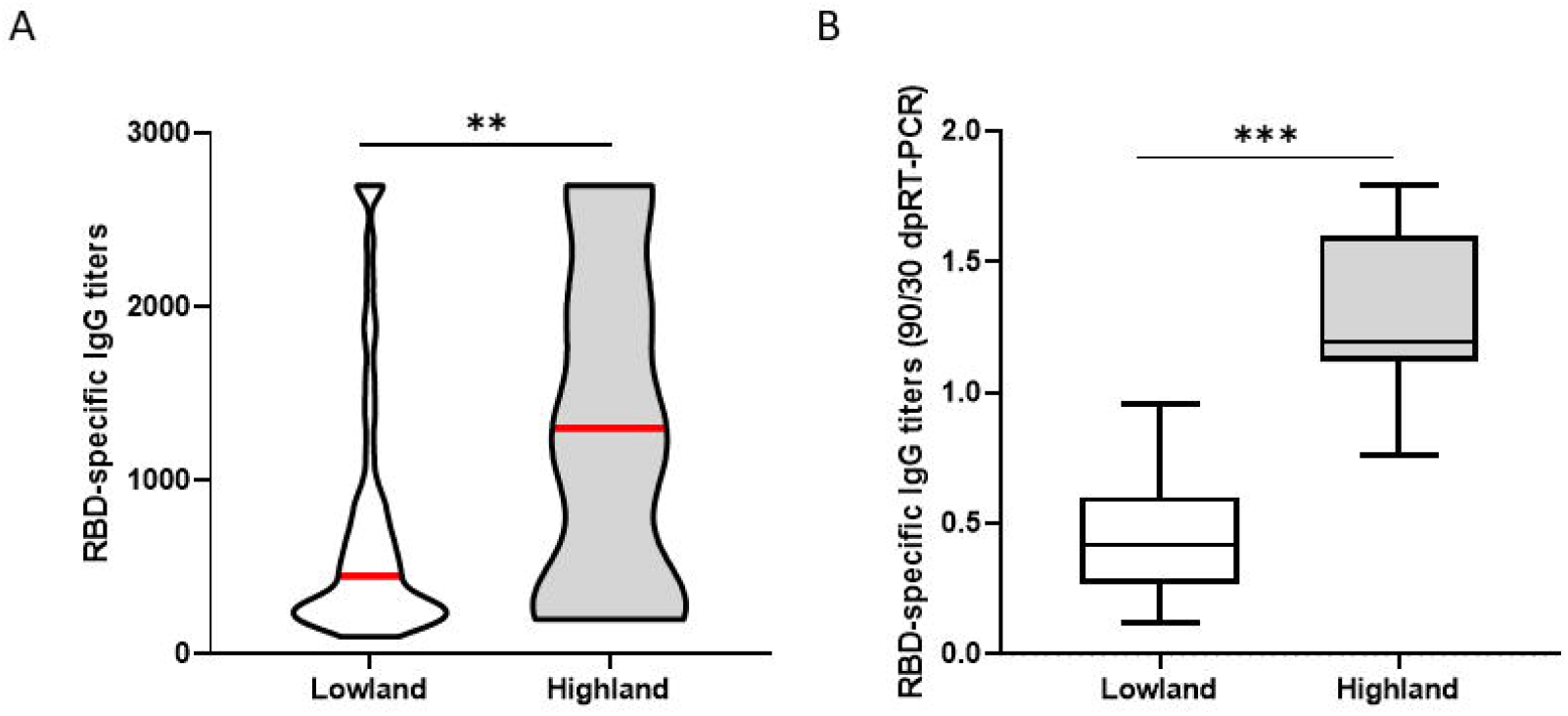
High and long lasting anti-RBD humoral response in a high altitude population. Populations at 431 mamsl (low altitude) and 2014 mamsl (high altitude) were assayed for the presence of IgG antibodies elicited against the RBD at 30 and 90 days post-diagnosis by RT-PCR for SARS-CoV-2. **A)** Specific IgG titers elicited at day 30 post-diagnosis in each population. Graph represent the mean ± S.E.M. **?p 0.001. **B)** Evolution of humoral immune response against SARS-CoV-2 after 90 days post-diagnosis. Results represent the ratio between RBD-specific IgG titers at day 90 and day 30 post-diagnosis.

## Discussion

The new coronavirus (SARS-CoV-2) infection has reached every continent, with new variants spreading quickly. As second and third waves of infections arise, and vaccination efforts attempt to ramp up, there is no consensus as to how long this pandemic could last. Among patients infected with SARS-CoV-2, the progression of disease is highly variable. Eighty percent of patients that become infected develop mild or no symptoms; whereas the remaining 20% develop moderate to severe disease ^35,36^. SARS-CoV-2 pathogenicity, results from an acute excessive virus replication followed by an uncontrolled inflammation and an exacerbated immunity. As the virus replicates, the adaptive immunity is stimulated to generate cellular and humoral responses in order to control the viral infection.

The role of sensitive molecular diagnostic techniques, such as RT-PCR and rapid antigen tests, are essential for the diagnosis of SARS-CoV-2 infection. Nevertheless, immunoserological tests have evolved as an indispensable tool, for example, in screening of potential plasma donors with high titers of anti-SARS-CoV-2 neutralizing antibodies, since the proven success of convalescent plasma therapy for COVID-19 ^4^. Many approaches have demonstrated that protection against SARS-CoV-2 is positively correlated with the development of high titers of neutralizing antibodies 17-20. Due to its role in viral entry to the host cell, the RBD of S emerged as a potential target antigen for the development of preventive and therapeutic strategies against COVID-19 ^20,37^. Equally as important is the usefulness of RBD for the diagnosis of SARS-Cov-2 infection, as supported by epidemiological data and molecular diagnosis ^33,34,38^. However, the unavailability of sensible, robust and cost-effective anti-RBD IgG diagnostic kit in our region motivated us to develop an “*In House*” ELISA for the detection of anti-RBD humoral responses against SARS-CoV-2.

By creating a stable cell line expressing high levels of RBD-His we were able to ensure a sufficient amount of antigen that enabled us to test thousands of patients from the public health system (SIPROSA-Tucumán, Argentina) (Fig 1). The immobilization of this antigen to ELISA plates allowed for the assembly of a highly sensitive and specific ELISA assay that showed an AUC of 0.991, for the detection of anti-RBD IgG elicited after SARS-CoV-2 infection. According to the traditional academic point system, an AUC between 0.90-1.0 indicates the antigen is an excellent ligand to correctly discriminate between the two groups (infected and non–infected) ^39^. The ELISA test developed did not show cross reactivity with pre-pandemic sera from patients infected with other common infections.

Importantly, the ability of the “*In House*” ELISA to correctly discriminate the occurrence or not of a SARS-CoV-2 infection was compared with other molecular and serological tests. We observed that RBD specific IgG antibodies were elicited in 60% of patients, previously diagnosed as SARS-CoV-2 positive. It has been reported that following infection, antibodies directed against RBD and N antigens begin to be detectable at slightly different times and in different amounts ^40,41^. Therefore, high levels of antibodies elicited against one antigen do not imply the presence of similar amounts of the other (Fig. 2). Nevertheless, our “*In House*” ELISA test showed high concordance with the commercial CMIA Architect by Abbott in discriminating presence or absence of IgG antibodies against SARS-CoV-2.

Our “*In House*” ELISA allowed surveying the immune response against SARS-CoV-2 RBD in patients of the public health system. Figure 4A revealed that 19% of individuals previously infected with SARS-CoV-2 displayed titers above 1350. According to Salazar et al. ^21^, these values correlate to a probability ≥80% of having neutralizing titers 160. Our data showed that one out of five plasma from COVID-19 recovered individuals were suitable candidates as donor for convalescent plasma therapy. Elevated anti-RBD IgG titers were found in patients above 60 years old compared to under 40 and 40-60 age groups. We also found that only 9.7% out of 196 female patients tested negative for anti-RBD IgG, compared to 48.9% in the case of males. This is in accordance with the fact that female sex is associated with greater SARS-CoV-2 antibody levels in disease early phase ^42^. However, the average titers induced by each sex group showed no statistical difference. Among the population studied, we noted that a high proportion of samples extracted from patients that reside in high altitude villages showed high anti-RBD IgG antibodies. Further analysis confirmed that patients from Tafí del Valle, a mountain village of 15000 residents situated at 2014 mamsl, presented increased levels of antibodies against RBD compared to patients from San Miguel de Tucumán, located at a much lower elevation (431 mamsl). To our knowledge, this is the first report that correlates high titers of anti-RBD antibodies with altitude. This suggests that a hypobaric hypoxia-adapted mechanism may play, at least in part, an important role in triggering a long-lasting humoral immune response of SARS-CoV-2, and might help explain previous publications reporting altitude as a protective factor for COVID-19 ^29 27,28^. These results provide an important quantitative target for therapeutic and prophylactic treatments.

## Data Availability

Data will be uploaded if required

## Acknowledges

This work was supported by grants from SkyBIO LLC, the School of Medicine of the UNT and Tucumán Goverment. We also acknowledge the partial support from Argentinean Research Council-CONICET (PIP 722 and 806), Argentinean Research Agency (PICT-MINCYT3379, and PICT2018-02989), Tucuman National University Grant (PIUNT-UNT D644/1 and D624). Diego Ploper was supported by Fundación Florencio Fiorini award We are particularly grateful to Dra. María Gabriela Simesen de Bielke, Graciela de Gorostiza from the Central Blood Bank of Tucumán (Banco Central de Sangre de Tucumán) for providing prepandemic serum samples. We are also thankful to Dr. Mateo Martinez and Dra. Roxana Toledo from the School of Medicine (UNT), and to Ing. Luis Rocha, Ing. Marina Gandur and Dr. Christian Jaroszewski from Medical Technology Department (SiProSa), Farm. Betina Heredia from Sterilization Department from Hospital del Niño Jesús (SiProSa), and CITED (Cámara de Instituciones Educ. Terap. Disc.) for their valuable contribution to the development of the present work.

## Abbreviations

RBD: Receptor binding domain
ROC: receiver operating characteristic curve
ELISA: Enzyme linked immunosorbent assay
mamsl: meters above median sea level

